# Cortico-thalamic tremor circuits and their associations with deep brain stimulation effects in essential tremor

**DOI:** 10.1101/2024.09.23.24314066

**Authors:** Shenghong He, Timothy O West, Fernando R Plazas, Laura Wehmeyer, Alek Pogosyan, Alceste Deli, Christoph Wiest, Damian M Herz, Thomas Simpson, Pablo Andrade, Fahd Baig, Michael G Hart, Francesca Morgante, James J. FitzGerald, Michael T Barbe, Veerle Visser-Vandewalle, Alexander L Green, Erlick A Pereira, Hayriye Cagnan, Huiling Tan

**Affiliations:** Medical Research Council Brain Network Dynamics Unit, University of Oxford, Oxford, UK; Nuffield Department of Clinical Neurosciences, University of Oxford, Oxford, UK; Department of Bioengineering, Imperial College London, London, UK; Department of Stereotactic and Functional Neurosurgery, University Hospital Cologne, and Faculty of Medicine, University of Cologne, Cologne, Germany; Neurosciences and Cell Biology Institute, Neuromodulation and Motor Control section, St. George’s, University of London, London, UK; Section of Movement Disorders and Neurostimulation, Department of Neurology, Focus Program Translational Neuroscience (FTN), University Medical Center of the Johannes Gutenberg-University Mainz, Mainz, Germany; Nuffield Department of Surgical Sciences, University of Oxford, Oxford, UK

**Keywords:** Essential tremor, deep brain stimulation, efferent and afferent, directed connectivity, local field potential

## Abstract

Essential tremor (ET) is one of the most common movement disorders in adults. Deep brain stimulation (DBS) of the ventralis intermediate nucleus (VIM) of the thalamus and/or the posterior subthalamic area (PSA) has been shown to provide significant tremor suppression in patients with ET, but with significant inter-patient variability and habituation to the stimulation. Several non-invasive neuromodulation techniques targeting other parts of the central nervous system, including cerebellar, motor cortex, or peripheral nerves, have also been developed for treating ET, but the clinical outcomes remain inconsistent. Existing studies suggest that pathology in ET may emerge from multiple cortical and subcortical areas, but its exact mechanisms remain unclear. By simultaneously capturing neural activities from motor cortices and thalami, and hand tremor signals recorded via accelerometers in fifteen human subjects who have undergone lead implantations for DBS, we systematically characterized the efferent and afferent cortico-thalamic tremor networks. Through the comparisons of these network characteristics and tremor amplitude between DBS OFF and ON conditions, we further investigated the associations between different tremor network characteristics and the magnitude of DBS effect. Our findings implicate the thalamus, specifically the contralateral hemisphere, as the primary generator of tremor in ET, with a significant contribution of the ipsilateral hemisphere as well. Although there is no direct correlation between the cortico-tremor connectivity and tremor power or reduced tremor by DBS, the strength of connectivity from the motor cortex to the thalamus and vice versa at tremor frequency predicts baseline tremor power and effect of DBS. Interestingly, there is no correlation between these two connectivity pathways themselves, suggesting that, independent of the subcortical pathway, the motor cortex appears to play a relatively distinct role, possibly mediated through an afferent/feedback loop in the propagation of tremor. DBS has a greater clinical effect in those with stronger cortico-thalamo-tremor connectivity involving the contralateral thalamus, which is also associated with bigger and more stable tremor measured with an accelerometer. Interestingly, stronger cross-hemisphere coupling between left and right thalami is associated with more unstable tremor. Together this study provides important insights into a better understanding of the cortico-thalamic tremor generating network and its implication for the development of patient-specific therapeutic approaches for ET.

## Introduction

Essential tremor (ET) is one of the most common movement disorders in adults, with an estimated prevalence of 0.5-5%.**^1-3^** Based on a series of cortico-cortical, cortico-muscular, and intermuscular coherence analyses, Raethjen and colleagues proposed that tremor in ET emerges from a number of cortical and subcortical motor centres, with each node acting as a dynamically changing oscillator and temporarily entraining each other.**^4-6^** In line with this theory, various neuromodulation techniques targeting distinct brain regions or other components of the central nervous system have been clinically or experimentally employed to treat ET. In clinical practice, high-frequency continuous deep brain stimulation (DBS) specifically targeting the Ventralis Intermediate Nucleus (VIM) of the thalamus has been widely employed and demonstrated significant efficacy in suppressing tremor in patients with ET. Additionally, alternative targets, such as the posterior subthalamic area (PSA, including zona incerta (ZI)), have also been proposed.**^7-11^** However, despite these promising clinical outcomes, notable inter-patient variability and habituation to the stimulation have been observed. In the realm of experimental non-invasive neuromodulation, several techniques have been developed for treating ET. This includes transcranial alternating/direct current stimulation (TACS/TDCS) targeting cerebellar**^12-14^** or motor cortex**^15^**, repetitive transcranial magnetic stimulation (rTMS) targeting cerebellar**^16-18^** or motor cortex**^19-20^**, and electrical stimulation targeting peripheral nerves**^21-22^**, although the clinical outcomes remain inconsistent. To optimize the efficacy of both invasive and non-invasive neuromodulatory approaches, a more precise understanding of the underlying mechanisms driving tremor in ET is needed. This entails elucidating the intricate interplay of multiple cortical and subcortical brain regions involved in the pathophysiology of ET.**^4-6^** However, most of the existing studies are only based on recordings from a single node in the motor circuit (cortical or subcortical) and lack within-subject pre- and post-intervention comparisons. Thus, the characteristics of cortical- and subcortico-tremor networks as well as how they change with intervention targeting the relevant nodes are still unclear.

In this study, based on the simultaneous recording of cortical EEG, thalamic local field potentials (LFPs), and limb acceleration measurements from patients with ET, we characterized cortico-thalamo-tremor networks through a directed connectivity analysis called generalized Orthogonalized Partial Directed Coherence (gOPDC),**^23^** and explored the associations between cortico-thalamo-tremor network characteristics and hand tremor characteristics. Furthermore, based on the data recorded during DBS OFF and DBS ON from each individual participant, we further investigated how the cortico-thalamo-tremor network characteristics predict DBS effect in tremor suppression.

## Materials and methods

### Human subjects and experimental protocol

Fifteen patients (mean age = 69.1 ± 7.26 years; mean disease duration = 21.1 ± 14.5 years; six females) with ET that underwent DBS surgery (30 DBS leads) participated in this study (P1-P7 and P12 were published previously).**^24^** All participants underwent bilateral implantations of DBS electrodes targeting the VIM thalamus and/or PSA/ZI area. The experimental protocol involved a posture holding task performed while sitting comfortably in a chair, with both arms raised up to the height of shoulders (**Fig. 1A**). The task was performed in blocks in both DBS OFF and ON conditions, with each block lasted about 20 s. There was a resting period when both arms were put down between two posture holding blocks (**Fig. 1B**). In average, the posture holding task was performed for 195.92 ± 11.54 s (mean ± SEM) and 196.67 ± 14.76 s in DBS OFF and ON conditions, respectively. The study was approved by the local ethics committees and all participants provided their informed written consent according to the Declaration of Helsinki. Clinical details of all participants are summarised in **Table I**.

**Figure 1.**
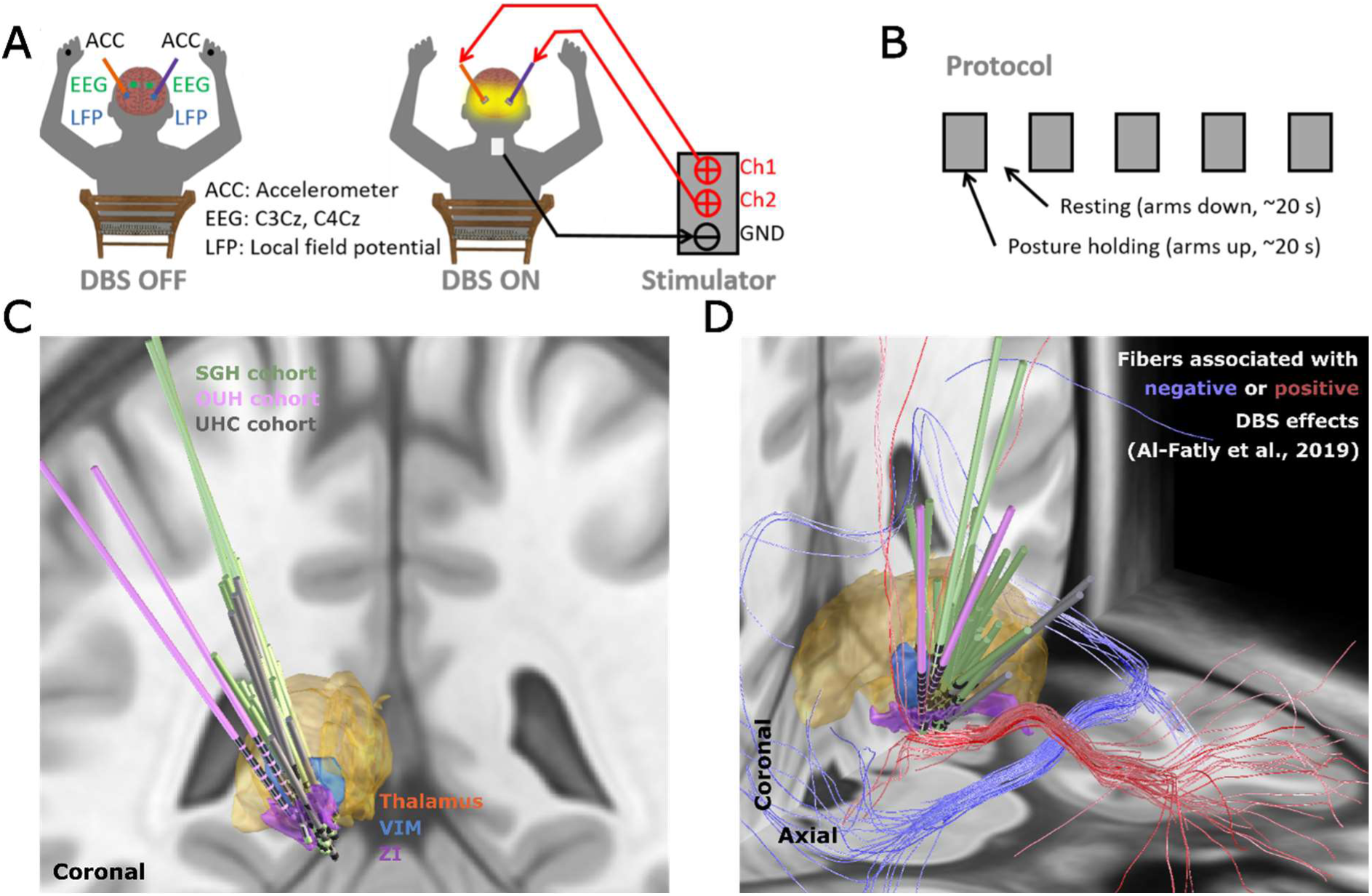
Experimental protocol. **(A)** Schematic of the posture holding task performed when the DBS is switched OFF (left) and ON (right). **(B)** Timeline for the experimental protocol which consists of 10 posture holding blocks (∼20 s per block) when both arms are raised up, and 10 resting blocks when both arms are put down. **(C)-(D)** 3D reconstruction in coronal (C) and coronal-axial (D) views of all analyzed DBS leads localized in standard Montreal Neurological Institute (MNI)-152_2009b space using Lead-DBS.**^25-26^** Electrodes in the left hemisphere were mirrored to the right hemisphere. UHC = University Hospital Cologne; OUH = Oxford University Hospital; SGH = St George’s Hospital; VIM = ventral intermediate thalamus; ZI = zona incerta.

**Table 1.**
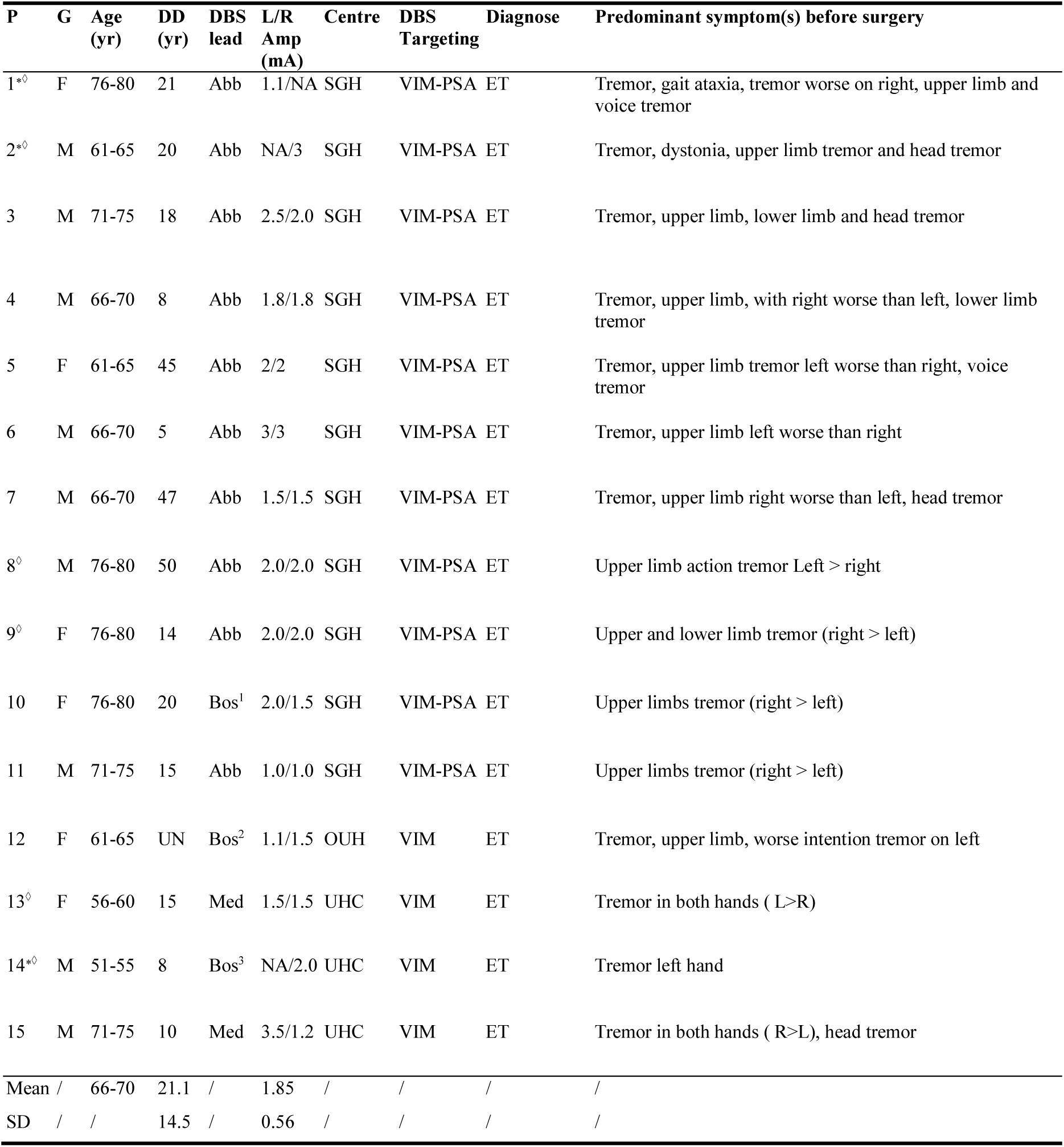
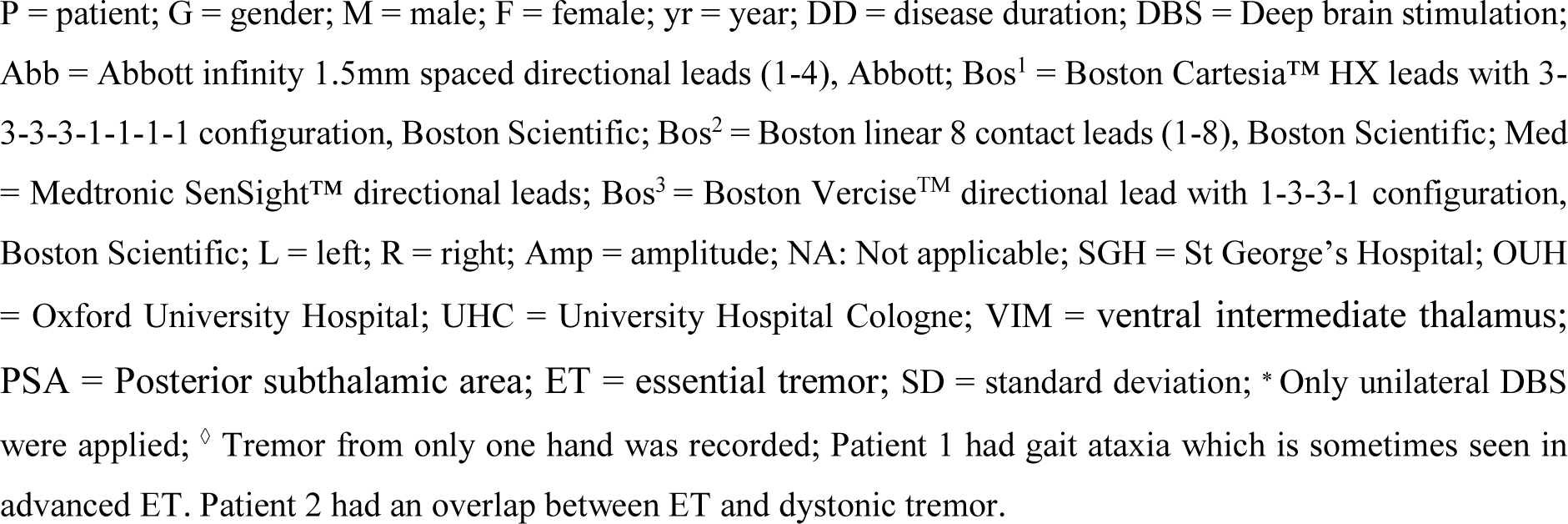
Clinical details of all recorded participants.

### Stimulation

Stimulation was applied bilaterally (except for P1, P2, and P14 who received unilateral stimulation contralateral to the tremor dominant hand) using a highly configurable custom-built neurostimulator or a CE marked stimulator (Inomed, Germany, or Bionics, Australia). In this study, monopolar stimulation was delivered with a fixed stimulation frequency of 130 Hz, a pulse width of 60 µs, and an interphase gap of 20 µs. These parameters are illustrated in **Supplementary Figure 1**. Here, the interphase gap is the time between the anodic and cathodic phases used in biphasic stimulation, previously shown to increase stimulation efficiency and reduce battery consumption.**^27-30^** The stimulation reference was connected to an electrode patch attached to the back of the participant (**Fig. 1A**). These stimulation parameters and configurations were selected based on previous literature.**^24,31-33^** The stimulation contact was selected as following: 1) contact levels targeting VIM-PSA area based on imaging data and/or feedback from neurosurgeon after operation were initially considered. 2) Among them, a contact searching procedure was applied to select the final stimulation contact for each hemisphere. Specifically, we delivered continuous DBS initially at 0.5 mA, then progressively increased the amplitude in 0.5 mA increments, until clinical benefit was seen without side effects such as paraesthesia, or until 3.5 mA was reached as the maximum amplitude. In average, the amplitude used in this study was 1.89 ± 0.12 mA (mean ± SEM). Details of the stimulation configuration for each participant are summarised in **Table I**.

### Data recording

Recordings from fifteen participants were conducted 1 to 5 days after the electrode implantation, when the DBS leads were temporarily externalized. While performing the posture holding task illustrated in **Fig. 1**, bilateral LFPs, EEGs covering “Cz”, “C3”, “C4”, “CPz”, “CP3”, and “CP4” according to the standard 10–20 system, and limb accelerations acquired using tri-axial accelerometers taped to the back of both hands were simultaneously recorded using a Porti (TMS International) amplifier at a sampling rate of 2048 Hz (for P1-P7, and P12), or a Saga amplifier (TMS International) at a sampling rate of 4096 Hz (for P8-P11, and P13-P15). When a Porti amplifier was used, the segmented contacts were first constructed in ring mode, then LFPs from two adjacent levels or two levels neighbouring the stimulation contact were recorded in the differential bipolar mode, to avoid saturation during stimulation. While LFPs from each individual contact were recorded in monopolar mode when a Saga amplifier was used, as it has a much higher tolerance of DC offset that may induce saturation during stimulation. Due to lack of tremor on the other hand after DBS surgery, limb accelerations were recorded only from one hand for six (P1-P2, P8-P9, and P13-P14) out of the 15 participants (**Table 1**), resulting in 24 tremulous upper limbs.

### Data analysis

#### Pre-processing

For the LFPs recorded in monopolar mode, bipolar signals were achieved offline by differentiating the recordings from two adjacent contacts or two contacts neighbouring the stimulation contact. In the cases with directional leads, only the contact pairs facing the same direction were considered. For the recorded EEGs, bipolar signals were constructed offline by differentiating between “C3” and “Cz” (i.e., “C3Cz”), or “C4” and “Cz” (i.e., “C4Cz”). The bipolar LFPs and EEGs as well as the recorded acceleration measurements were band-pass filtered at 1–95 Hz and then band-stop filtered at 48-52 Hz using two 4^th^ order zero-phase Butterworth IIR digital filters in MATLAB (R2023-b, MathWorks). After filtering, a principal component analysis (PCA) was applied on the tri-axial acceleration measurements, and the first component was selected as the measurement of tremor on a given hand. PCA components reflect a linear combination of the three (orthogonal) axes, with the first component reflecting the orientation that captures the maximum variance in the data. This technique has precedence in previous studies.**^13,34^** To consider the natural intra-individual tremor variability during posture holding (**Fig. 2A**), we split the data into non-overlapping 2 s segments and considered each segment as a trial. This procedure resulted in 98.0 ± 5.8 (mean ± SEM) and 98.3 ± 7.4 trials per subject in DBS OFF and DBS ON conditions, respectively.

**Figure 2.**
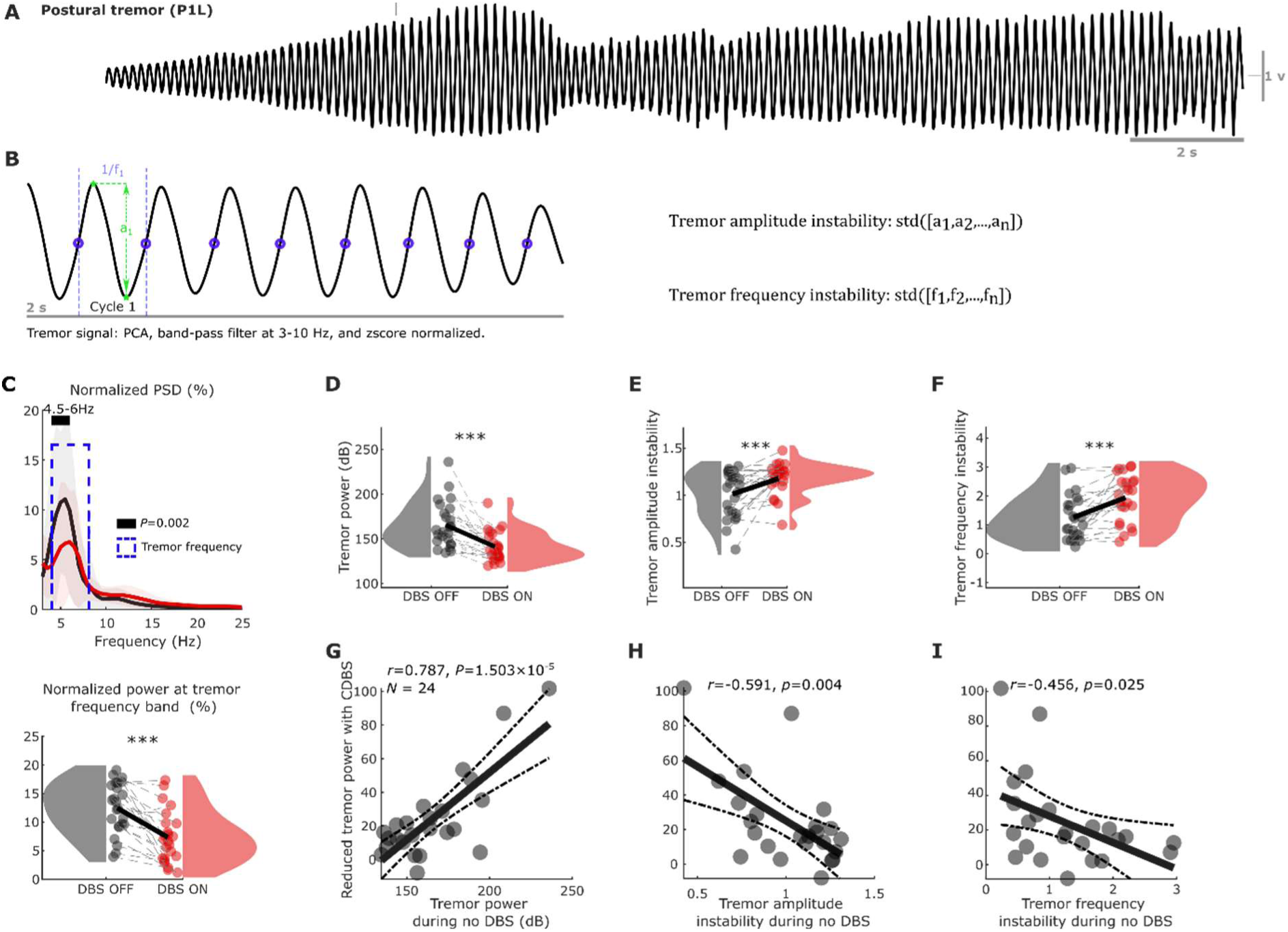
Comparisons of tremor characteristics between DBS OFF and DBS ON conditions. **(A)** An example of 30-s postural tremor (P1L) showing the instability of tremor in ET. **(B)** Demonstration of the quantifications of tremor amplitude and frequency instability from a segment of 2 s measurement from an accelerometer. **(C)** Normalized power spectral density (PSD) of accelerometer measurements showed peaks at tremor frequency band in both DBS OFF (black) and DBS ON (red) conditions (upper panel), with a significant reduction of the normalized power (in percentage) in the individualized tremor frequency band during DBS ON (lower panel). **(D)-(F)** Comparisons of tremor power (D), amplitude instability (E), and frequency instability (F) between DBS OFF (black) and DBS ON (red) conditions using raincloud plots.**^51^**Here the shaded areas indicate distributions (probability density) of the data. **(G)-(I)** Tremor power during DBS OFF (baseline) positively (G) while tremor amplitude (H) and frequency (I) instability negatively correlated with the reduction in tremor power during DBS (Pearson correlation). Solid lines in C and bars in C-F indicate mean, while shaded areas in C and error bars in C-F indicate standard error of the mean (SEM). Statistics were applied between DBS OFF and DBS ON conditions using a nonparametric cluster-based permutation procedure in C (PSD) on a hand-by-hand basis, or using generalized linear mixed effect modelling in all bar plots (C-F) on a trial-by-trial basis. Multiple comparisons were corrected by controlling the false discovery rate (FDR). *** P < 0.001 after FDR correction.

#### Spectral analysis

After pre-processing, power spectral density (PSD) was estimated using Welch’s overlapped segment averaging estimator for each individual LFPs, EEGs, and acceleration measurements in each trial,**^35^** in a frequency range of 1 to 95 Hz with a 0.5 Hz resolution. To select the tremor frequency for each hand in each trial, we first normalized the PSD of the acceleration measurement against the sum of the power between 1 and 25 Hz, then the frequency between 3 and 10 Hz that has the maximum power was selected as the tremor frequency. To select one bipolar LFP for each hemisphere, we averaged the normalized PSD across trials for each bipolar LFP channel, and selected the one with maximum power at the averaged tremor frequency of both tremor hands. Furthermore, for each trial (i.e., 2-s segment), the normalized PSD and power (raw and normalized) at the tremor frequency were calculated for EEGs (“C3Cz” and “C4Cz”), acceleration measurements (left and right hand), and the selected bipolar LFPs for further analysis.

#### Tremor instability analysis

After pre-processing, tremor amplitude and frequency instability in each trial were quantified for each hand. Specifically, the acceleration measurements were high- and low-pass filtered at 3 and 10 Hz using two sixth order zero-phase Butterworth IIR digital filters, and z-score normalized. Then, zero-crossing points from negative to positive were used to identify individual tremor cycle within each trial. For each tremor cycle, the instantaneous tremor amplitude was quantified as the distance between the peak and trough, while instantaneous tremor frequency was defined as the reciprocal of the duration of the tremor cycle, as shown in **Fig. 2B**. Finally, tremor amplitude and frequency instability were quantified as the standard deviation of the instantaneous tremor amplitude and frequency across cycles. Please note that with z-score normalization, these represent how stable the tremor is in terms of amplitude and frequency within the 2-s segment, as demonstrated in Supplementary Fig. 2. Tremor stability index**^13,34^** and multiscale entropy (MSE)**^36^** have previously been proposed to distinguish ET and parkinsonian tremor. Thus these measurements were also computed for comparison.

#### Connectivity analysis

Based on the simultaneously recorded cortical, subcortical, and tremor signals, we investigated the cortico-thalamo-tremor network characteristics through a directional connectivity analysis using a method called generalized orthogonalized partial directed coherence (gOPDC) developed by Omidvarnia et al.**^23,37^** In this method, signal power was first orthogonalized before quantifying coherence, to mitigate the effect of volume conduction.**^38^** Briefly, a coefficient of a multivariate autoregressive (MVAR) model was converted to the spectral domain using the Fourier transform, and then used to calculate the power spectral density matrix. Prior to frequency domain conversation, the MVAR coefficients were orthogonalized.**^37^** This effectively minimizes shared variance between the autoregressive components of the signals, such that correlations arise from off-diagonal terms (i.e., connectivity). Further details can be found in Omidvarnia et al.**^23,37^** Only the imaginary part of the orthogonalized partial directed coherence (OPDC) was considered to reduce spurious correlations introduced by factors such as movement/tremor artefact. In addition, the scale invariant version of the classical PDC (i.e., gOPDC) was used to handle numerical problems associated with different variance of signal amplitudes in LFPs, EEGs, and acceleration measurements (known as time-series scaling).**^39-40^** This method has been shown to reliably detect event-related directional information flow at ∼10 Hz based on non-overlapping 1-s segments of neonatal EEGs.**^23^** In the current study, we are mainly interested in the tremor frequency band at 3-8 Hz thus the data was truncated into 2-s non-overlapping segments. Based on gOPDC, the mean efferent (from cortices/thalamus to tremor) and afferent (from tremor back to cortices/thalamus) connectivity in a frequency range covering 2 Hz around the basic tremor frequency as well as 2 Hz around the second harmonic frequency were analysed. Furthermore, direct and indirect causal effects of a certain structure were explored by comparing the unconditioned versus conditioned gOPDC models, i.e., excluding or including the corresponding source.**^23^** Each gOPDC measurement was compared against its surrogate distribution. To this end, the pre-processed continuous tremor time-series was divided into two segments according to a randomly selected point (with a minimum of 2 s margin on each side) and then swapped back and forth to disrupt the coupling between EEG/LFP and tremor signals. Then, the shuffled data were truncated into non-overlapping 2 s trials. This procedure was repeated until we got 1000 trials of shuffled data. The same gOPDC metrics were derived from the shuffled data, resulting in a surrogate distribution of 1000 values per measurement.**^41^** This approach ensured that any signatures of connectivity remaining, following disruption of the EEG/LFP and tremor signal pairs, arose from the independent statistics of each signal.

#### Spatial distributions of the connectivity measurements

Lead placements were confirmed by fusion of preoperative MRI and postoperative CT scans, which were further established by reconstructing the electrode trajectories and location of different contacts using the Lead-DBS MATLAB toolbox (version 2.6.0).**^25^** The electrode locations were registered and normalized into the Montreal Neurologic Institute (MNI) 152-2009b space using the Connectomic ET Target Atlas.**^11^** As shown in Fig. 1C and D, most of the tested electrodes targeted the VIM-PSA area, close to the fibers, suggested to provide positive DBS effects in tremor patients.**^11^** To investigate the spatial distributions of the bidirectional gOPDC connectivity (thalamo-cortical and cortico-thalamic) and their associations with different targets for ET, we repeated the connectivity analyses for all available bipolar LFP channels from all patients, and mapped them onto the MNI space based on the coordinates of each contact. In addition, for each hemisphere, the volume of tissue activated (VTA) during stimulation was estimated using a finite element method (FEM),**^25^** based on the individual electrode position used for the connectivity calculation and a common stimulation amplitude (i.e., 1 mA). Subsequently, the intersections between the VTA and different subcortical structures (e.g., VIM and ZI) were quantified and used to correlate with different connectivity measurements.

### Statistical analysis

Statistical analyses were conducted using custom-written scripts in MATLAB R2023-b (The MathWorks Inc, Nantucket, MA).

To compare the PSD of EEGs, LFPs, and acceleration measurements between DBS OFF and DBS ON conditions, a non-parametric cluster-based permutation procedure (repeated 2000 times) was applied, in which multiple comparisons were controlled theoretically.**^42^**

To compare the tremor characteristics (power, amplitude instability, and frequency instability) or gOPDC measurements quantified on a trial-by-trial basis between different conditions (e.g., DBS OFF versus DBS ON, unconditioned versus conditioned gOPDC models, or real gOPDC versus its null distribution), generalized linear mixed effect (GLME) modelling was used.**^43-44^** We also used GLME to further investigate the associations between gOPDC measurements and tremor characteristics on a trial-by-trial basis. In each GLME model, the slope(s) between the predictor(s) and the dependent variable were set to be fixed across all tremor hands while a random intercept was set to vary by hand. The parameters were estimated based on maximum-likelihood using Laplace approximation, the Akaike information criterion (AIC), estimated value with standard error of the coefficient (*k* ± SE), multiple comparisons corrected *P*-value and proportion of variability in the response explained by the fitted model (*R*^2^) were reported. Here multiple comparisons applied to different measurements were corrected using false discovery rate (FDR) approach.**^45-46^**

To explore the correlations between different tremor characteristics or gOPDC measurements and the effect of DBS in tremor suppression, or between different gOPDC measurements, Pearson correlation was applied on a hand-by-hand basis. For each correlation analysis, the pairwise linear correlation coefficient (*r*), multiple comparisons corrected *P*-value (based on FDR), and sample size (*N*) were reported. Here the sample size was equal to the number of tremulous upper limbs (*N*=24), unless outliers were identified according to the Pauta criterion (3*σ* criterion).

## Results

### 1. Continuous DBS reduces tremor power and stability, and the DBS effect correlates with baseline tremor power and instability

The amplitude of postural tremor in ET is unstable over time,**^47-50^** as shown in **Fig. 2A**, which motivated us to quantify tremor characteristics including power at tremor frequencies (peak frequency ± 1 Hz), tremor amplitude instability, and frequency instability in non-overlapping 2 s epochs, as shown in **Fig. 2B** (with more details in Methods). As expected, there was a significant reduction in tremor power during DBS ON compared with DBS OFF (**Fig. 2C**, PSD at 4.5-6 Hz: t = 3.799, P = 0.002; normalized power at individualized tremor frequency band: k = -5.280 ± 0.120, P < 1 × 10^-4^; **Fig. 2D**, absolute power at individualized tremor frequency band: k = -26.502 ± 0.621, P < 1 × 10^-4^), although tremor-frequency peaks were identified in both DBS OFF and DBS ON conditions. This was accompanied by a significant power reduction at the tremor frequency band in the VIM thalamic LFPs (**Supplementary Fig. 3A and B**) and cortical EEGs (**Supplementary Fig. 3C and D**). In addition, DBS significantly increased the instabilities of tremor amplitude (**Fig. 2E**, k = 0.173 ± 0.011, P < 1 × 10^-4^) and frequency (**Fig. 2F**, k = 0.744 ± 0.029, P < 1 × 10^-4^). Here k indicates estimated value with standard error of the coefficient using generalized linear mixed effect (GLME) modelling (Methods). Apart from an expected positive correlation between the level of tremor reduction with DBS and the baseline tremor power during DBS OFF (**Fig. 2G**, r = 0.787, P = 1.50 × 10^-5^), baseline tremor instability was also found to be negatively correlated with the effect of DBS (**Fig. 2H**, amplitude instability, r = -0.591, P = 0.004; **Fig. 2I**, frequency instability, r = -0.456, P = 0.025). We repeated this analysis using two other tremor instability measurements including TSI**^13,34^** and MSE**^36^**. As shown in **Supplementary Fig. 4**, these measurements were highly correlated with each other and showed similar relationships with respect to the effect of DBS. Together, these suggested that more severe and stable tremor during DBS OFF was associated with a larger effect of DBS on tremor reduction.

### 2. The efferent and afferent thalamic-tremor networks are both lateralized and interact across hemispheres

Based on the simultaneously recorded hand acceleration measurements and bilateral thalamic LFPs during posture holding (**Fig. 3A**), we characterized bidirectional connectivity between VIM thalamus and hand tremor in the tremor frequency band (2 Hz around the peak tremor frequency as well as 2 Hz around the second harmonic frequency) using generalized Orthogonalized Partial Directed Coherence (gOPDC, with details in Methods). As shown in **Supplementary Table 1**, we first tested the main effects of laterality (contralateral versus ipsilateral), cross-hemisphere coupling (conditioned versus unconditioned), and directionality (efferent versus afferent), as well as the interaction effects between them. This analysis revealed significant main effects for all these conditions and significant interaction effects between laterality and directionality, as well as between cross-hemisphere coupling and directionality. We then conducted pairwise comparisons and the results revealed that without DBS, the efferent connectivity from the contralateral thalamus to hand tremor was significantly stronger than that from the ipsilateral thalamus (**Fig. 3C**, unconditioned model, k = -0.001 ± 0.001, P = 0.029; hemisphere conditioned model, k = -0.001 ± 0.001, P = 0.011), as expected. However, the afferent network showed an opposite pattern, with a significantly stronger input from hand tremor to the ipsilateral thalamus than that to the contralateral thalamus (**Fig. 3D**, unconditioned model, k = 0.002 ± 0.001, P = 0.001; hemisphere conditioned model, k = 0.003 ± 0.001, P = 4.73 × 10^-5^). Overall, the strength of the afferent network was stronger than the efferent network. This thalamic-tremor network laterality disappeared during DBS (**Supplementary Fig. 5**). Compared with the model only involving unilateral (either contralateral or ipsilateral) thalamus and hand tremor (**Fig. 3B left**, unconditioned model), conditioning the impact from the other thalamus (hemisphere conditioned model, **Fig. 3B right**) significantly reduced the efferent connectivity from both the contralateral (**Fig. 3C**, k = -0.002 ± 0.001, P = 0.004) and ipsilateral (**Fig. 3C**, k = -0.002 ± 0.001, P = 0.002) thalami to hand tremor. Similarly, the afferent connectivity from hand tremor to both the contralateral (**Fig. 3D**, k = -0.004 ± 0.001, P = 7.88 × 10^-11^) and ipsilateral (**Fig. 3D**, k = -0.004 ± 0.001, P = 2.91 × 10^-8^) thalami were also significantly reduced in the hemisphere conditioned model compared with unconditioned model. This suggests that there was cross-hemisphere coupling between the two thalami in the thalamic-tremor network. During DBS, the hemisphere conditioned model also significantly reduced the efferent connectivity from both thalami to hand tremor, but not the afferent connectivity from hand tremor to both thalami (**Supplementary Fig. 5**). The details of the GLME models used for these tests were summarized in **Supplementary Table 1**.

**Figure 3.**
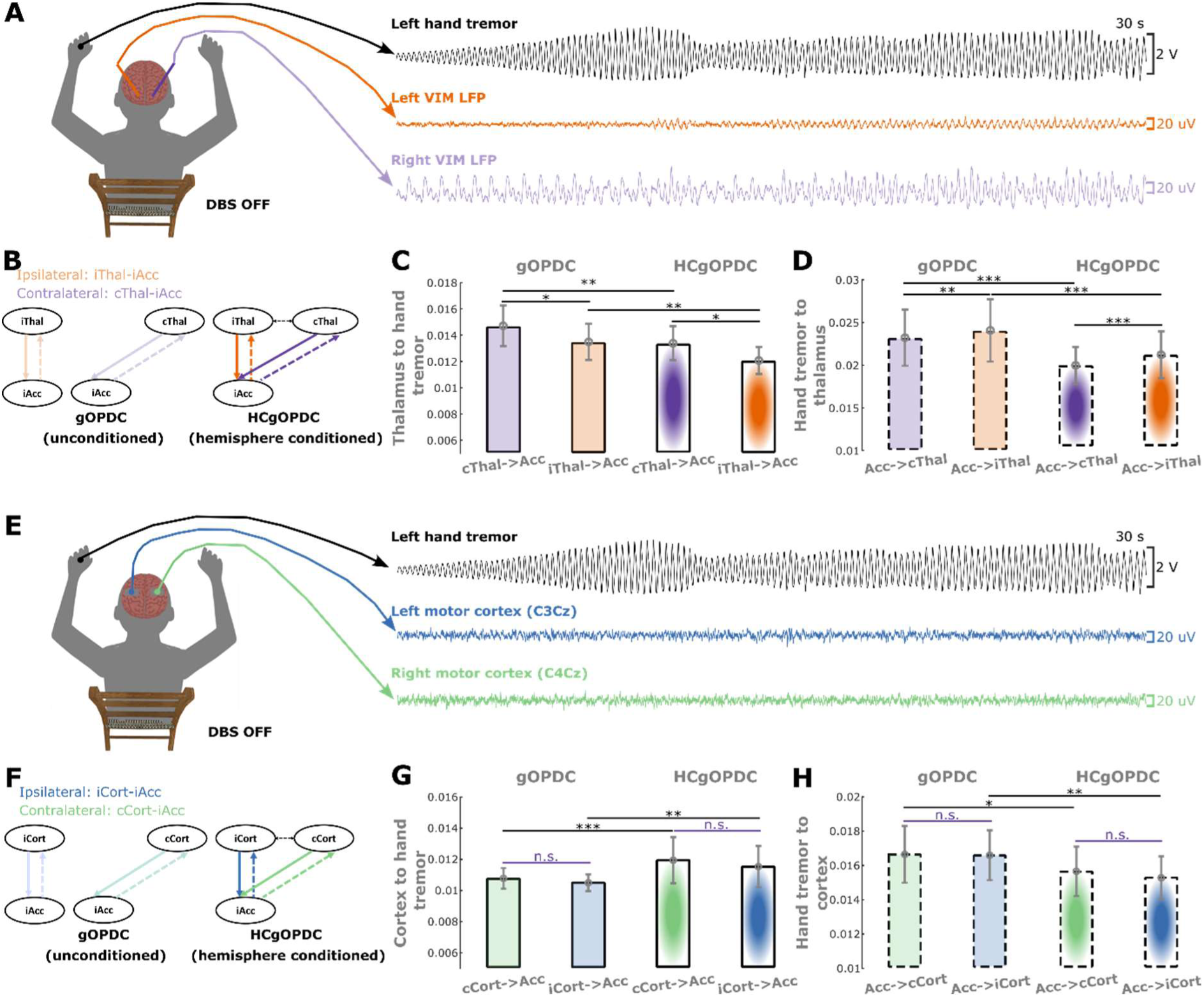
Characteristics of thalamic-tremor and cortico-tremor networks when DBS was switched off. **(A)** A demonstration of left-hand postural tremor and thalamic LFP recordings from participant 1, left hand (P1L) during DBS OFF condition. **(B)** Directed connectivity between VIM thalamus and hand tremor quantified using generalized Orthogonalized Patial Directed Coherence (gOPDC). Solid lines indicate efferent connectivity from thalamus to hand tremor, while dashed lines indicate afferent connectivity from hand tremor to thalamus. Orange and purple represent the connectivity with ipsilateral and contralateral VIM thalami, respectively. The upper and lower panels indicate gOPDC involving only one thalamus (unconditioned) and both thalami (hemisphere conditioned: HCgOPDC), respectively. **(C)** Efferent connectivity from the contralateral thalamus was significantly stronger than that from the ipsilateral hemisphere in both unconditioned (left) and hemisphere conditioned (right) models. When conditioning the impact from the other hemisphere, the efferent connectivity from the contralateral (purple) and ipsilateral (orange) thalami to hand tremor were both significantly reduced. **(D)** Afferent connectivity from hand tremor to the contralateral thalamus was significantly weaker than that to the ipsilateral hemisphere in both unconditioned (left) and hemisphere conditioned (right) models. When conditioning the impact from the other hemisphere, the afferent connectivity from hand tremor to the contralateral (purple) and ipsilateral (orange) thalami were both significantly reduced. **(E)-(H)** The same as (A)-(D) but for cortico-tremor network. Bars and error bars indicate mean and standard error of the mean (SEM), respectively. Statistics were applied on each comparison using generalized linear mixed effect modelling on a trial-by-trial basis. Multiple comparisons were corrected by controlling the false discovery rate (FDR). * P < 0.05; ** P < 0.01; *** P < 0.001; after FDR correction.

### 3. The efferent and afferent cortico-tremor networks are non-lateralized but interact across hemispheres

Based on the simultaneously recorded hand acceleration measurements and EEGs from bilateral sensorimotor cortices during posture holding (**Fig. 3E**), we characterized bidirectional (efferent and afferent) connectivity between cortical activities and hand tremor in the tremor frequency band using gOPDC. Similarly, we first identified significant main effects on cross-hemisphere coupling and directionality, but not on laterality. The interaction between cross-hemisphere coupling and directionality was also significant (Supplementary Table 2). We then conducted pairwise comparisons and the results. We then conducted pairwise comparisons and the results revealed that without DBS, there was no significant difference between the efferent connectivity from the contralateral and ipsilateral motor cortices to hand tremor in either the unconditioned (**Fig. 3G**) or hemisphere-conditioned model. Similar results were observed in the afferent tremor to cortical connectivity (**Fig. 3H**). Compared with the model only involving unilateral sensorimotor cortex and hand tremor (**Fig. 3F left**, unconditioned model), conditioning the impact from the other cortex (conditioned model, **Fig. 3F right**) significantly increased the efferent connectivity from both the contralateral (**Fig. 3G**, k = 0.001 ± 4 × 10^-4^, P = 9.0 × 10^-4^) and ipsilateral (**Fig. 3G**, k = 0.001 ± 4 × 10^-4^, P = 0.003) sensorimotor cortices to hand tremor. However, the afferent connectivity from hand tremor to both the contralateral (**Fig. 3H**, k = -0.001 ± 0.001, P = 0.030) and ipsilateral (**Fig. 3H**, k = -0.001 ± 4 × 10^-4^, P = 0.007) cortices reduced significantly in the conditioned model compared with unconditioned model. During DBS, none of these comparisons were significant (**Supplementary Fig. 6**). These results suggest that the cortico-tremor network is not lateralized but interacts across hemispheres, in other words, there is coupling between the ipsilateral and contralateral cortices, and both of them contribute to hand tremor equally. The details of the GLME models used for these tests were summarized in **Supplementary Table 2**.

### 4. Interaction between the thalamic-tremor and cortico-tremor networks

To investigate the potential relationship between the thalamic-tremor and cortico-tremor networks, we included the simultaneously recorded hand tremor, bilateral thalamic LFPs, and cortical EEGs in a single gOPDC model (network conditioned gOPDC: NCgOPDC). By comparing the efferent connectivity strength achieved from this network conditioned model (**Fig. 4A**) against those achieved from the gOPDC model only involving thalamic (**Fig. 3B**) or cortical (**Fig. 3E**) sources, we found that when conditioning the cortical inputs, the efferent connectivity from thalamus to hand tremor was significantly reduced (**Fig. 4B**, DBS OFF, k = -0.002 ± 0.001, P = 8.75 × 10^-4^; DBS ON, k = -0.002 ± 0.001, P = 9.25 × 10^-6^). Vice versa, conditioning thalamic inputs significantly reduced the efferent connectivity from cortex to hand tremor (**Fig. 4C**, DBS OFF, k = -0.003 ± 0.001, P = 3.57 × 10^-7^; DBS ON, k = -0.002 ± 0.001, P = 2.35 × 10^-6^). Similarly, the afferent connectivity from hand tremor to thalamus (**Fig. 4E**, DBS OFF, k = -0.004 ± 0.001, P = 5.60 × 10^-6^; DBS ON, k = -0.002 ± 0.001, P = 5.05 × 10^-5^) or cortex (**Fig. 4F**, DBS OFF, k = -0.006 ± 0.001, P < 1 × 10^-4^; DBS ON, k = -0.002 ± 0.001, P = 2.67 × 10^-4^) in the network conditioned model (**Fig. 4D**) was also significantly reduced compared with the gOPDC model only involving thalamic (**Fig. 3B**) or cortical (**Fig. 3E**) sources. These results suggest that the thalamic-tremor and cortico-tremor networks interact with each other, in line with the theory proposed by Raethjen et al.**^4-6^** When directly comparing the connectivity from thalamus to cortex versus the connectivity from cortex to thalamus (**Fig. 4G**), we found that the connectivity from cortex to thalamus was significantly stronger than the connectivity in the other direction (from thalamus to cortex, **Fig. 4H**). The results were similar for either tremor (k = 0.005 ± 0.001, P = 3.60 × 10^-17^), alpha (k = 0.007 ± 0.001, P = 9.89 × 10^-29^), or beta (k = 0.004 ± 4 × 10^-4^, P = 9.59 × 10^-23^) frequency bands.

**Figure 4.**
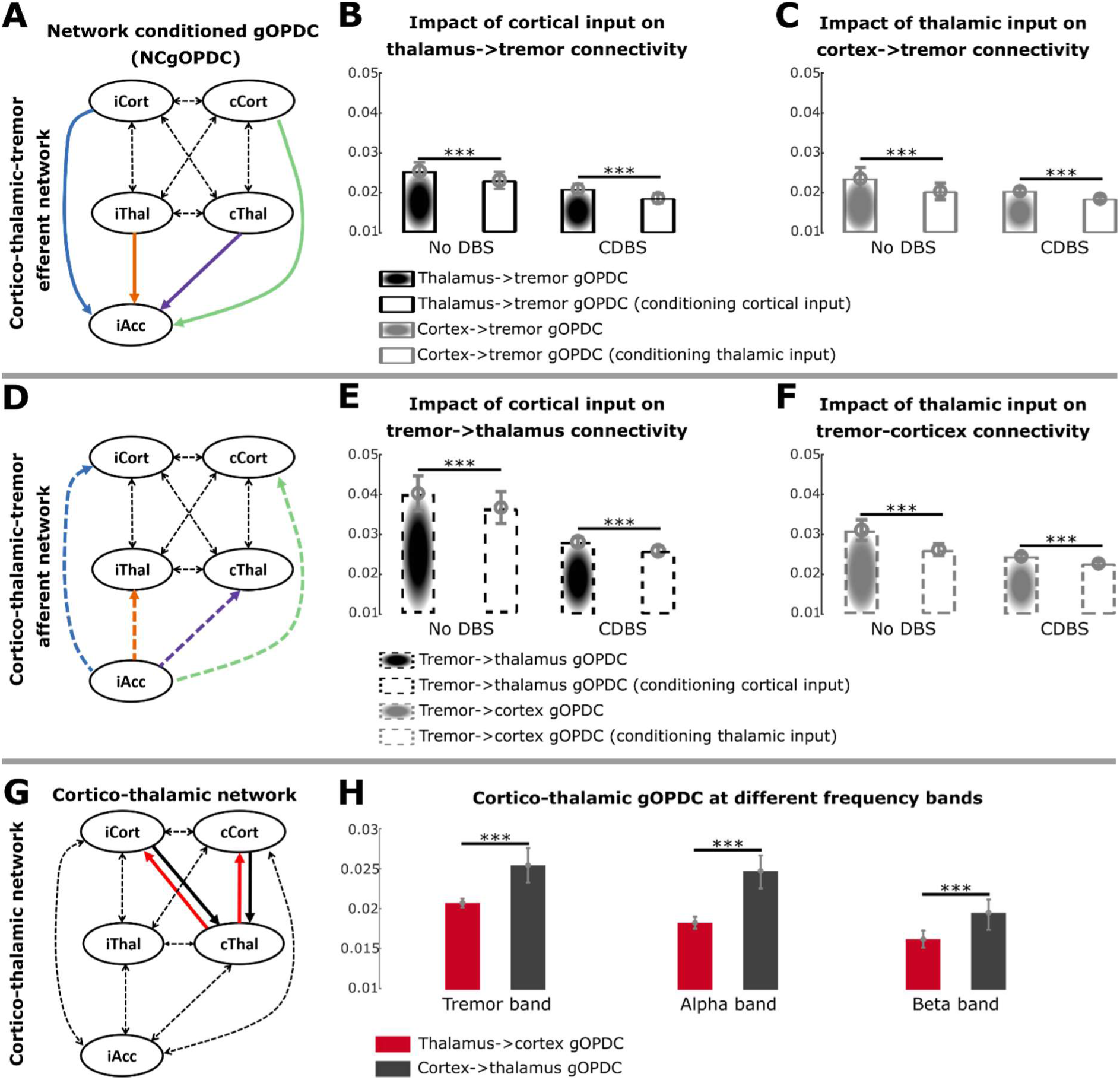
Characteristics of cortico-thalamo-tremor network. **(A)** Directed efferent connectivity from sensorimotor cortex and VIM thalamus to hand tremor quantified using generalized Orthogonalized Patial Directed Coherence (gOPDC). **(B)** Comparing with the model only involving bilateral thalami in Fig. 3, conditioning cortical input significantly reduced the efferent connectivity from thalamus to hand tremor in both DBS OFF and DBS ON conditions. **(C)** Comparing with the model only involving bilateral sensorimotor cortices in Fig. 3, conditioning thalamic input significantly reduced the efferent connectivity from cortex to hand tremor in both DBS OFF and DBS ON conditions. **(D)** Directed afferent connectivity from hand tremor to sensorimotor cortex and VIM thalamus quantified using gOPDC. **(E)** Comparing with the model only involving bilateral thalami in Fig. 3, conditioning cortical input significantly reduced the afferent connectivity from hand tremor to thalamus in both DBS OFF and DBS ON conditions. **(F)** Comparing with the model only involving bilateral sensorimotor cortices in Fig. 3, conditioning thalamic input significantly reduced the afferent connectivity from hand tremor to cortex in both DBS OFF and DBS ON conditions. Here the connectivity in (A)-(F) was quantified in tremor frequency band. **(G)** Directed connectivity between sensorimotor cortices and the contralateral VIM thalamus relative to the focused hand tremor quantified using gOPDC. **(H)** The directed top-down connectivity from cortex to thalamus (black) was significantly and consistently stronger than bottom-up connectivity from thalamus to cortex (red) in tremor (left), alpha (middle), and beta (right) frequency bands. Bars and error bars indicate mean and standard error of the mean (SEM), respectively. Statistics were applied on each comparison using generalized linear mixed effect modelling on a trial-by-trial basis. Multiple comparisons were corrected by controlling the false discovery rate (FDR). *** P < 0.001 after FDR correction.

### 5. Connectivity involving contralateral thalamus positively correlates with DBS effect

To further investigate whether the cortico-thalamo-tremor network characteristics could be used to predict the effect on tremor suppression with VIM DBS, we performed Pearson’s correlation analysis between different connectivity measurements and the DBS effect in reducing tremor. This analysis revealed that the efferent connectivity from the contralateral thalamus to hand tremor (**Fig. 5A**, r = 0.54, P = 0.017) and the overall connectivity strength between thalamus and cortex at tremor frequency (thalamus to cortex plus cortex to thalamus, **Fig. 5C**, r = 0.556, P = 0.017) positively correlated with the level of tremor power reduction during DBS ON. There was a trend of positive correlation between the efferent connectivity from the ipsilateral thalamus and hand tremor, which however did not survive multiple comparison correction (**Fig. 5B**, r = 0.431, P = 0.071). Combining all connectivity involving the contralateral thalamus increased the effect size of the positive correlation (**Fig. 5D**, r = 0.617, P = 0.014). In addition, there was no correlation between the reduced tremor power and the efferent connectivity from either the contralateral (**Fig. 5E**) or ipsilateral (**Fig. 5F**) sensorimotor cortex, or the overall connectivity strength between thalamus and cortex in other frequency bands as control (**Fig. 5G**, alpha band; **Fig. 5H**, beta band). When using generalized linear mixed effect modelling (GLME) to predict tremor power using various connectivity measurements (**Supplementary Table 3** Model 1), only the connectivity involving thalamus including efferent connectivity from contralateral (k = 94.488 ± 21.8, P = 4.571 × 10^-5^) and ipsilateral (k = 116.54 ± 24.651, P = 1.44 × 10^-5^) thalami to hand tremor, connectivity from thalamus to cortex (k = 88.322 ± 22.94, P = 2 × 10^-4^), and connectivity from cortex to thalamus (k = 41.844 ± 16.178, P = 0.015) in tremor frequency band showed significant prediction effects, but not the efferent connectivity from sensorimotor cortex to hand tremor. To test if the connectivity measurements are simply representations of electrode locations. We quantified the distances between the selected contacts and a sweetspot in VIM for tremor suppression with DBS suggested in a previous study,**^11^** and correlated them with connectivity measurements and DBS effects. The results showed that the connectivity measurements in **Fig. 5A-D** did not correlate with the distances between contacts and the tremor sweetspot (**Supplementary Fig. 7A-D**), but provided better prediction of DBS effects than the distances (**Supplementary Fig. 7E**).

**Figure 5.**
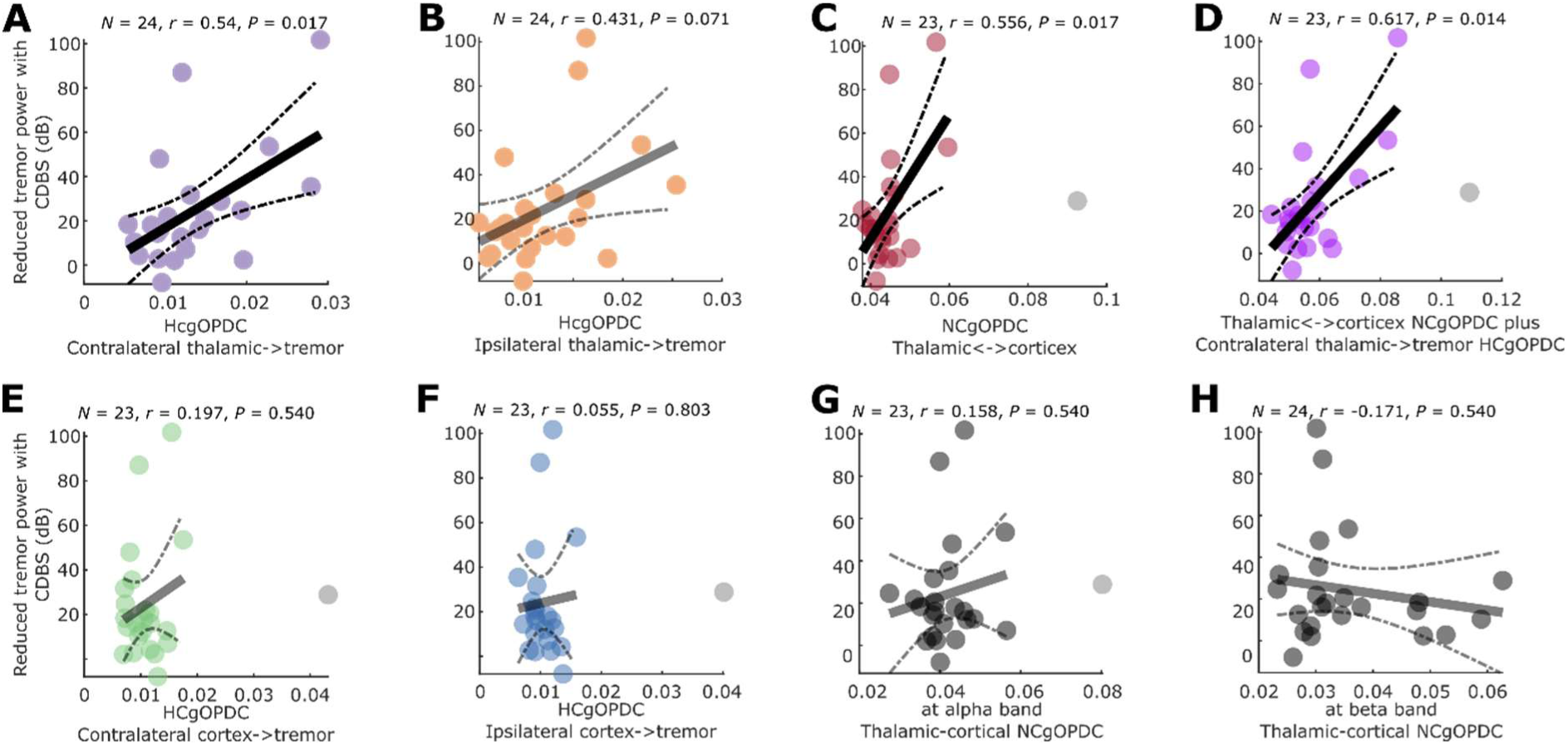
Correlations between cortico-thalamo-tremor network characteristics and the reduced tremor power with DBS. **(A)-(B)** Correlations between the efferent connectivity from the contralateral (A) or ipsilateral (B) thalami to hand tremor and the reduced tremor power with DBS. **(C)** Correlation between the sum of thalamus to cortex and cortex to thalamus connectivity at tremor frequency band and the reduced tremor power with DBS. (D) Correlation between the sum of all connectivity at tremor frequency involving the contralateral thalamus and the reduced tremor power with DBS. **(E)-(F)** There was no correlation between the efferent connectivity from the contralateral (E) or ipsilateral (F) sensorimotor cortices to hand tremor and the reduced tremor power with DBS. **(G)-(H)** There was no correlation between the sum of thalamus to cortex and cortex to thalamus connectivity at alpha (G) or beta (H) frequency band and the reduced tremor power with DBS. P-values were corrected for multiple comparisons by controlling false discovery rate (FDR).

### 6. Thalamic-tremor connectivity is predicted by tremor characteristics

We then used GLME to test if the thalamic-tremor connectivity strength can be predicted by tremor characteristics (power and instability). This analysis revealed that stronger tremor power (**Supplementary Table 3 Model 2**, k = 0.0002 ± 3.88 × 10^-5^, P = 9.12 × 10^-8^) and smaller tremor amplitude instability (indicating more stable tremor, **Supplementary Table 3 Model 2**, k = -0.007 ± 0.002, P = 0.001) together predicted greater connectivity involving contralateral thalamus. On the other hand, stronger tremor power (**Supplementary Table 3 Model 3**, k = -0.001 ± 4 × 10^-4^, P < 1 × 10^-4^) and greater connectivity involving the contralateral thalamus (**Supplementary Table 3 Model 3**, k = -0.685 ± 0.236, P = 0.004) together predicted smaller tremor amplitude instability, i.e., more stable hand tremor. These results confirmed that there is a clear association between the strength of the functional connectivity involving the contralateral thalamus and tremor characteristics.

### 7. Motor cortex and thalamus have separate pathways in tremor propagation

Although the thalamo-cortical and cortico-thalamic connectivity at tremor frequency predicted the DBS effects (**Fig. 5C and D**), there was no correlation between them (**Fig. 6A**). In addition, the strongest thalamo-cortical connectivity and cortico-thalamic connectivity clustered at different areas in the MNI space (**Fig. 6B and C**). These results suggested that the thalamo-cortical and cortico-thalamic connectivity at tremor frequency band may have different spatial sources. Using Lead-DBS, we quantified the VTA during stimulation at 1 mA for each hemisphere, as shown in **Fig. 6D**. Correlation analysis revealed that the intersection between VTA and VIM thalamus positively correlated with the thalamo-cortical connectivity (**Fig. 6E**, r = 0.38, P = 0.038), but not the cortico-thalamic connectivity (r = 0.03, P = 0.452) measured from the same contacts. In contrast, the intersection between VTA and ZI positively correlated with the cortico-thalamic connectivity (**Fig. 6F**, r = 0.50, P = 0.021), but not the thalamo-cortical connectivity (r = 0.12, P = 0.274). The results were consistent when using 2 mA amplitude for simulation in Lead-DBS. Together, these results suggest that tremor propagation from thalamus to motor cortex mainly involves VIM, while propagation from the motor cortex back to thalamus mainly involves ZI/PSA.

**Figure 6.**
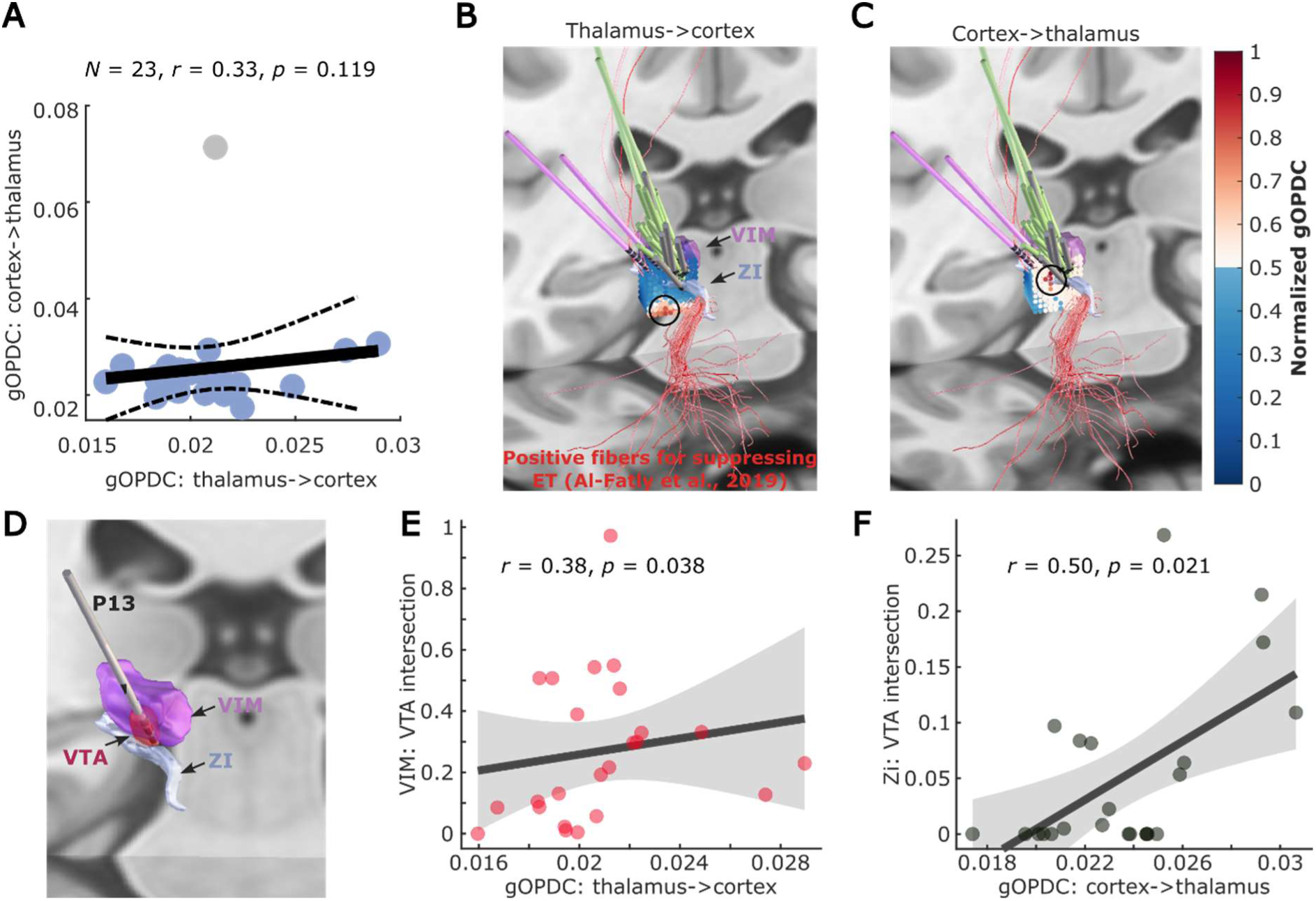
Comparisons between thalamo-cortical and cortico-thalamic connectivity. **(A)** Directed connectivity at tremor frequency band (gOPDC) from thalamus to cortex (x-axis) did not correlate with that from cortex to thalamus (y-axis). **(B)-(C)** The strongest thalamo-cortical (B) and cortico-thalamic (C) gOPDC clustered at different areas in the standard MNI-152_2009b space. **(D)** A demonstration of the volume of tissue activated (VTA) with DBS at 1 mA applied to the selected bipolar LFP channels (P13). **(E)** Results from Spearman rand correlation between the intersection of the VTA in VIM thalamus and directed connectivity from thalamus to cortex. **(F)** Results from Spearman rand correlation between the intersection of the VTA in ZI and directed connectivity from cortex to thalamus.

## Discussion

In this study, we characterized the cortico-thalamo-tremor network based on hand acceleration measurements, thalamic LFPs, and cortical EEGs recorded simultaneously from people with ET during posture holding in both ON and OFF DBS conditions **(Fig. 7)**. Specifically, we have shown that apart from with a stronger lateralized efferent connectivity from the contralateral thalamus to hand tremor (as expected), there is also contribution from the ipsilateral thalamus, as evidenced by significant changes observed in connectivity measurements between thalamus and hand tremor (efferent and afferent) when partializing out (conditioning) the contribution made by the ipsilateral thalamus. The lateral asymmetry was not observed in the cortico-tremor network. Furthermore, although the thalamic-tremor and cortico-tremor networks have different network characteristics and correlated differently with tremor, they interact with each other, with significant changes observed in the connectivity when partializing out (conditioning) the contribution made by the other network. Secondly, we have shown that both the tremor power during DBS OFF and the effect of VIM/PSA DBS were only predicted by the connectivity involving the thalamus but not by the cortico-tremor connectivity. In addition, the connectivity involving the contralateral thalamus, which showed the best correlation with the DBS effect, was independently predicted by tremor power and amplitude instability, suggesting both tremor power and tremor instability represent some level of underlying cortico-thalamo-tremor network characteristics. Lastly, although both thalamo-cortical and cortico-thalamic connectivity at tremor frequency band contributed to predicting DBS effect on tremor suppression, there was no correlation between them, suggesting motor cortex and thalamus may have separate pathways in tremor propagation. These results together shed light on the tremor network in ET.

**Figure 7.**
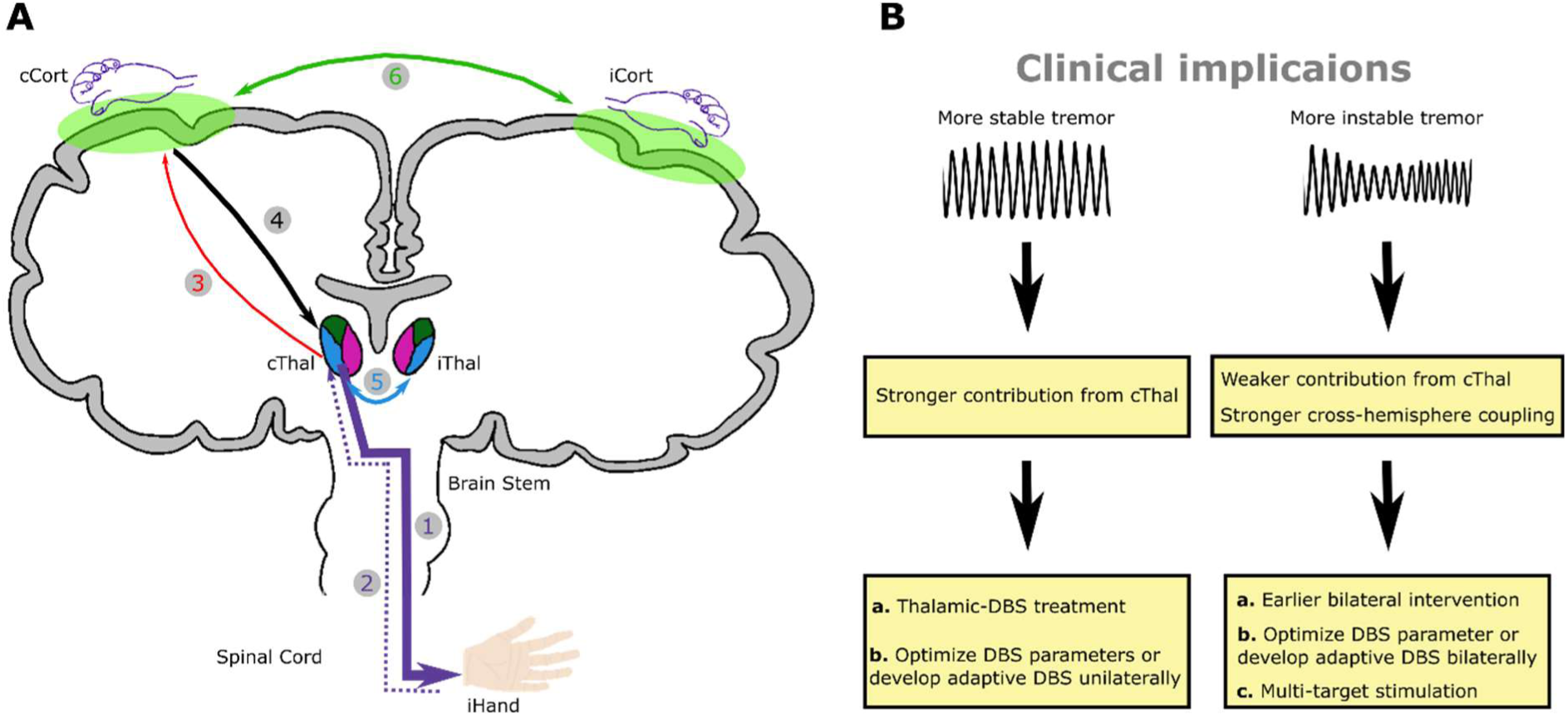
A summary of the current study. **(A)** Our study suggests that tremor in ET originates from the contralateral thalamus (path 1). The motor cortex is involved through an indirect pathway, likely via a feedback loop, by receiving afferent input from the tremulous hand through ascending pathways (paths 2 and 3) and sending it back to the thalamus (path 4). There is also significant cross hemisphere-coupling at both subcortical (path 5) and cortical (path 6) levels. **(B)** Potential clinical implications of this study. cCort=contralateral motor cortex; iCort=ipsilateral motor cortex; cThal=contralateral thalamus; iThal=ipsilateral thalamus.

### Verification of the gOPDC connectivity measurements

In this study, the tremor information flow was assessed using partial directed coherence, quantified using a method called gOPDC, which has been suggested to be able to remove common components akin to volume conduction effects. This method addresses the numerical problem associated with different variance of signal amplitudes (here accelerometer measurement, LFP, and EEG), and can detect directed information flow within a subsecond time scale in nonstationary multichannel signals.**^23^** A variant algorithm of this method (without orthogonalization) has also been used to characterize the cerebello-cortical network between essential, Parkinsonian, and mimicked tremor.**^52^** Results of a few tests provide evidence that the quantified gOPDC measurements are physiologically meaningful: 1) along with the reduction of tremor power during DBS, gOPDC measurements were significantly reduced with DBS compared with during DBS OFF (**Supplementary Table 4**), and the laterality of the thalamic-tremor network also disappeared (**Supplementary Fig. 5**); 2) We applied gOPDC to surrogate data by shuffling the tremor measurements relative to LFPs and EEGs. Statistical analysis showed that gOPDC measurements based on real data were all significantly bigger than those derived from surrogate data (**Supplementary Fig. 8** and Method); 3) The presented results were still valid when using the variant algorithm without orthogonalization (i.e., gPDC), which resulted in significantly lager connectivity values but has weaker effect sizes in the thalamic laterality and correlation analysis (**Supplementary Fig. 9**). Please note that the presented thalamic-tremor network laterality phenomenon was not captured by another non-directional connectivity measurement, i.e., imaginary coherence, in which the directionality (i.e., afferent and efferent) and causality are not considered (**Supplementary Fig. 10**).

### The contralateral thalamus as a main generator of tremor in ET

Existing studies showed that the tremor in ET remains constant when the resonant frequency of the oscillating limb is changed by added inertia.**^53-54^** Compared with Parkinsonian tremor, tremor in ET has a much narrower frequency tolerance (a measure that characterizes the temporal evolution of tremor by quantifying the range of frequencies over which the tremor may be considered stable), suggesting it has a more finely tuned central drive.**^13,55-56^** Thalamic neuronal activity correlated with ET.**^57^** Our results showed that only the thalamus-involved connectivity significantly correlated with both the tremor power during DBS OFF and the reduced tremor power during DBS ON, but not the cortico-tremor connectivity strength. Within the central thalamic-tremor network, the efferent connectivity from the contralateral thalamus to hand tremor was significantly stronger than that from the ipsilateral thalamus. This laterality was not due to the selection of analysed bipolar LFP channels, as it persisted when averaging across all bipolar LFP channels within each hemisphere (**Supplementary Fig. 11**). These results are consistent with existing literature showing strong coherence between thalamic LFP and contralateral muscular EMG in ET,**^57^** and clinical evidence demonstrating substantial tremor suppression in the contralateral hand following unilateral thalamic DBS.**^58-59^** This evidence suggests that the tremor might originally be generated from the contralateral thalamus. Whaley et al. reported that from a clinical series of 487 consecutive individuals diagnosed with ET, only about half (52%) of the sample reported bilateral initial tremor onset, but eventually about 90% of the individuals presented bilateral tremor.**^60^** Here we also found that there was a significant bidirectional cross hemisphere coupling within the thalamic-tremor network, highlighted by the significant changes in the efferent and afferent information flow between the contralateral/ipsilateral thalamus and accelerometer when partializing out the contributions from bilateral information flow (**Fig. 3C and D**). To further investigate if this is physiologically meaningful, we repeated the GLME modelling (Supplementary Table 3) by adding the gOPDC measurements between hemispheres in the models. The results showed that stronger cross-hemisphere communication predicted larger (e.g., power) but more unstable tremor (e.g., larger amplitude and frequency instability) (**Supplementary Table 5**). In addition, the afferent connectivity from hand tremor back to the ipsilateral thalamus was significantly stronger than that to the contralateral thalamus. However, this was only true for the selected bipolar LFP channels but not when averaging across all bipolar channels within each hemisphere (**Supplementary Fig. 11**). Together these results suggest that the ipsilateral thalamus still plays an important role in the development of tremor. Please note that effects of laterality, cross-hemisphere coupling, and correlations between thalamic-tremor connectivity and DBS effects were not driven by the fact that most of the patients included in this study presented bilateral dysfunction: our key results were not impacted when partializing out (conditioning) the contribution made by the other tremulous hand (Supplementary Fig. 12).

### Cortical involvement in ET

Conflicting results have been reported on the existence of tremor-related cortical activity in ET.**^61-62^** Raethjen et al. reported an intermittent loss of corticomuscular coherence at tremor frequency despite strong peripheral tremor constantly present.**^6^** Roy et al. showed that providing high visual feedback worsened tremor compared with low feedback.**^63^** Here we found the strength of the bidirectional cortico-thalamic connectivity predicted baseline tremor power during DBS OFF (Supplementary **Table 3**, Model 1) as well as the effect of DBS (**Fig. 5C**). Conditioning either the cortical or thalamic inputs significantly reduced the thalamic-tremor or cortico-tremor connectivity. These results support the presence of cortical involvement in tremor propagation in ET. In addition, we found that the afferent connectivity from hand tremor back to cortex negatively correlated with that to thalamus (Supplementary **Table 3**, Model 4), and the connectivity from cortex to thalamus was significantly stronger than the connectivity from thalamus to cortex, with no clear correlation between them (Supplementary **Table 3**, Model 5; **Fig. 6A**). Furthermore, we quantified cortico-thalamic and thalamo-cortical gOPDC at the tremor frequency band for each individual bipolar LFP channel for all recorded hemispheres, and mapped the values into standard MNI space using the Lead-DBS toolbox. This revealed the strongest cortico-thalamic and thalamo-cortical gOPDC clustered at relatively different areas relative to VIM thalamus, with both close to the fibers suggested to be associated with positive DBS effect in ET (**Fig. 6B-C**).**^11^** Furthermore, simulation analysis revealed that the intersection between the VTA and VIM thalamus correlated with thalamo-cortical gOPDC, but not cortico-thalamic gOPDC. In comparison, the intersection between the VTA and ZI correlated with cortico-thalamic gOPDC, but not thalamo-cortical gOPDC (**Fig. 6D-F**). There was, however, no correlation between the efferent cortico-tremor connectivity and tremor power or reduced tremor by DBS. Based on these results, we speculate that the cortical involvement in tremor propagation may primarily reflect sensory inputs from the muscles, relayed via ascending tracts like the dorsal column–medial lemniscus (DCML) pathway, incorporating the spinal cord and sensory thalamic areas. This process appears relatively independent from the cerebellar outflow pathways, involving the VIM-PSA region, which is likely more directly involved in tremor generation and is also a common target for DBS in the treatment of ET.**^52,64-65^** Further exploration on this would require new data and is outside the scope of this work.

### Clinical implications

Our results showed that thalamic-tremor connectivity correlated with the DBS effect on tremor suppression (**Fig. 5**). Linear mixed effect modelling revealed that both tremor power and tremor amplitude instability had independent contributions when predicting the directed connectivity involving the contralateral thalamus: more stable tremors associated with greater connectivity involving the thalamus, which predicted a greater DBS effect. This is consistent with previous studies showing that those with more stable tremors benefited more from tremor phase-specific DBS targeting the thalamus,**^66-67^** or phase-specific transcranial electrical stimulation targeting the cerebellum.**^14^** Our results also highlighted that more unstable tremor was associated with stronger cross-hemisphere coupling. The outcome of DBS in people with ET is heterogeneous with some patients not benefitting from the intervention or developing habituation over time. Lead placement may account for some of this heterogeneity in clinical outcomes. However another important factor to consider is that the clinical syndrome of ET might be underlined by different network characteristics. Indeed, these potential variations in the disease network may necessitate the use of alternative targeting and stimulation modalities. The following clinical implications arise from our study (**Fig. 7**). *1) Where to stimulate?* Thalamic DBS may be more effective for individuals with larger, more stable tremors since tremors with these characteristics are potentially driven by a more prominent tremor-generating source in the contralateral thalamus. On the other hand, our results suggest that unstable tremor arises from a less focal source and is more likely to involve multiple generators including those in the cortex. This may suggest that more unstable tremors may benefit from alternative surgical targets, such as the PSA or stimulation of multiple regions across the cerebello-thalamo-cortical pathway,**^11,68-69^** similar to the strategy that is currently being investigated in chronic pain, involving implantation of electrodes encompassing multiple targets to disrupt the pain-network rather than perturbing a single node.**^70-71^** *2) How to stimulate?* Our results show that patients with unstable tremors exhibit stronger cross-hemisphere coupling. This suggests that implanting DBS bilaterally may be more beneficial in these patients, even in the case that tremor may only initially present in one hand. Moreover, when assessing the effects of DBS on a tremulous hand, optimizing stimulation parameters on both sides may be more beneficial than focusing solely on the contralateral side. *3) When to stimulate?* Taking into account the variations in the disease network may also be beneficial for the development of a fully embedded closed-loop stimulation system. For instance, for those with more stable tremors, it might be more practical to implement closed-loop stimulation based on the thalamic LFPs.**^24^** While for those with more unstable tremors, additional sites might be needed for closed-loop stimulation.**^72^**

### Limitations

There are several limitations in the current study. First, all recordings were conducted 1-6 days after the first surgery of DBS electrode implantations, thus some participants might still experience an appreciable postoperative stun effect, which however is more likely to overall reduce rather than increase the effect size of the reported results. Second, although the associations between tremor and tremor network characteristics were explored on a trial-by-trial basis, the correlations between these characteristics and the effect of DBS were only investigated on a hemisphere basis, due to the lack of data to effectively quantify the reduced tremor in a trial-by-trial basis. Third, although we somehow characterized both thalamic-tremor and cortico-tremor networks, only a thalamus-targeted intervention was applied in this study, thus it is still unclear whether the cortico-tremor network characteristics could be used to predict the effect of cortex-targeted brain stimulation. Furthermore, although tests against surrogate distributions and comparisons between DBS OFF and ON conditions suggest that the cortico-tremor connectivity, quantified based on scalp EEG, is physiologically meaningful, it should be interpreted carefully and the use of intracranial cortical recordings such as electrocorticography (ECoG) should be preferred wherever possible to improve anatomical precision. Finally, we show that the thalamic-tremor network presented both laterality and cross-hemisphere dependency characteristics, but we cannot further investigate the potential of using these characteristics to predict the effect of unilateral DBS, as bilateral stimulation was applied for most of the patients in this study.

## Data availability

The data and codes will be shared on the data sharing platform of the MRC Brain Network Dynamics Unit: https://data.mrc.ox.ac.uk/mrcbndu/data-sets/search.

## Acknowledgement

This work was supported by the Medical Research Council (MC_UU_00003/2) and the Guarantors of Brain. S.H. was also supported by Royal Society Sino-British Fellowship Trust (IES\R3\213123). We thank all participants for making this study possible, thank Dr Bassam Al-Fatly and Dr Amir Omidvarnia for providing useful discussions on data analysis.

## Notes

### Competing Interest Statement

The authors have declared no competing interest.

### Author Declarations

Medical Sciences Interdivisional Research Ethics Committee of University of Oxford gave ethical approval for this work (REC No: 18/SC/0436).

